# The Smoldering Pandemic: Self-Reported Prevalence Assessment of Prolonged Grief Disorder. A cross sectional study of bereaving adults during the Covid Pandemic in Pakistan

**DOI:** 10.1101/2023.01.10.23284300

**Authors:** Ayesha Siddiqua, Shaaf Ahmad, Iqra Nawaz, Muhammad Zeeshan, Amina Rao

## Abstract

**Introduction:** The Covid-19 pandemic brought forward unprecedented psycho-social challenges for the world. The devastating loss of human lives created a burden of grief throughout the world. The bereaved were put at a greater risk of grief complications with high death tolls, strict social isolation guidelines and a halt to communal funeral practices. Prolonged Grief Disorder is a young psychiatric condition which refers to an abnormal grief reaction that exceeds the normal cultural, social and religious norms. In this study, we assessed the prevalence of Prolonged Grief Disorder (PGD), as mentioned in ICD-11 in Pakistan, along with its correlation to anxiety, depression and psychological distress. Severity of grief reactions were compared with the place of death and relationship with the deceased.

**Methods:** A cross sectional online survey was conducted during the month of October 2021. Sample size was calculated using OpenEpi and data was collected through non probability sampling. The questionnaire was validated and shared through multiple social forums. A total of 737 participants residing in Lahore Pakistan, who had lost a close one due to Covid-19 participated in the study. Demographics, loss related information, and self-reported symptoms measured by 13-item Prolonged Grief Disorder Scale, Patient Health Questionnaire-4 and Kessler-6 scales were obtained.

**Results:** The prevalence of Prolonged Grief Disorder was found to be 15.4%. There was a significant correlation of grief intensity with depression and anxiety.Prolonged Grief Disorder puts individuals at greater risk of suffering from serious mental illnesses. People who were closely related to the deceased were more likely to experience severe Prolonged Grief Disorder symptoms.

**Conclusion:** Early detection and treatment of high risk individuals is necessary to mitigate the burden of grief and associated risk of anxiety and depression. Overall we conclude that discussions pertaining to grief and measures to curb the psychological effects are crucial in the post-pandemic world.

## INTRODUCTION

The World Health Organization reported that COVID-19 has resulted in more than 4.5 million deaths around the world (1). In March 2020, three months after the first case was detected in Wuhan, China, the organization officially declared the viral outbreak as a ‘global pandemic. Isolation protocols and social distancing measures were put in place to curb the increasing cases. With all the challenges that the pandemic brought with it, the most difficult were the social and psychological repercussions that came during it; as it has been demonstrated that for every death by COVID-19, Nie and more bereaving family members were left behind (2). Bereavement is the term used to reflect the fact of having lost someone to death. Grief is used to describe the emotional, cognitive, functional, and behavioral responses to the death. Mourning is also sometimes used interchangeably with bereavement and grief, usually referring more specifically to the behavioral manifestations of grief, which are influenced by social and cultural rituals, such as funerals, visitations, or other customs (3).

With unexpected casualties, separation of families from patients, and absence of final goodbyes in hospitals ICUs (4, 5), traditional mourning practices were put to halt. Such conditions put the grieving family members at a greater risk of enduring psychological distress and developing prolonged grief disorder (PGD). The risk factors for developing PGD include low perceived social support (6), fewer years of education, depression, anxiety, (7) and experiencing death of a partner/child (8). As social animals, humans often rely on social support and emotional help from their family members, however, due to social distancing measures and bans on travel, bereaving individuals were left alone, at the disposal of their own selves, which contributed immensely to the development of PGD among them (9, 10).

Grief is not a state, rather a process which transitions over time from acute (intensely painful) to integrated, (whereby the reality of death is assimilated and the bereaved is able to engage in activities of life again). This process continues to evolve. There may be periods when acute grief reawakens, and in some individuals it may become intense and chronic.

Multiple terms and criteria have been used to define grief and the various transitions that it takes. It is described as Persistent Complex Bereavement Disorder (PCBD) in the Diagnostics and Statistical Manual of Mental Disorders (DSM-V) (11). The World Health Organization describes it as, Prolonged Grief Disorder (PGD) has now been added in the International Classification of Diseases-11 (ICD-11) for the first time. The ICD-11 Prolonged Grief Disorder is defined as a disturbance in which, following the death of a partner, parent, child, or other person close to the bereaved, there is persistent and pervasive grief response characterized by longing for the deceased or persistent preoccupation with the deceased accompanied by either by intense emotional pain.

Thus, studies estimating the prevalence and incidence of PGD among various throughout the world assume more importance. While there is a noted variance in the prevalence rates of PCBD and PGD (12), a longitudinal study shows that the difference between PCBD and PGD is only semantic (13).

PGD has been studied in multiple international studies (6, 14, 15) and among refugees (16, 17) as well. However, there is considerable heterogeneity in the prevalence of PGD depending upon the measurement tool employed. A meta-analysis, estimating prevalence rate of PGD in the adult bereaved population, demonstrated that 1 in 10 bereaving adults are at risk of developing PGD (18). In the context of a low-middle-income country (LMIC), such as Pakistan where there is limited scientific literature on the prevalence of grief and its complications, there is greater emphasis on communal funeral practices and mourning. As majority of the individuals identify as Muslims, not being able to perform all the religious rituals of funeral during the pandemic caused great grievance to families of the deceased. Thus, estimating the prevalence of PGD assumes greater importance among low- and middle-income countries.

The primary objective of this study is to assess the prevalence of PGD, among the bereaving adult population of Lahore, Pakistan, who had lost a family member/partner to COVID-19. It also aims to facilitate further research, early interventions, and awareness of this psychiatric condition as it has been reported that early detection of people with elevated bereavement distress and offering them preventive interventions (19) are effective in reducing the burden of PGD and fostering adaptations to loss (8) Our work also aims to investigate the sociodemographic factors associated with PGD, loss related factors, and association of severity of grief symptoms with depression, anxiety and psychological distress. Although PGD criteria given by WHO has increased international clinical applicability, (20) and better captures psychopathology (21), however, for diagnostic validity it requires structured clinical interviews (22), and newer self-reporting measures with cultural criteria scoring, which are in process of development (14).

## METHODS

A cross sectional study was conducted through a web-based survey form. Before entering the survey form a consent page included a brief introduction to the study, confidentiality statement, time required, and voluntariness of participation. Only when the participants ticked the box “I understand the information provided above and consent to be a part of this research”, then they completed the formal survey. Participants had the right to withdraw from the survey at any time without stating reason. Public grief support and counseling resources were linked at the start and end of the survey form. Ethical approval was obtained from the ethics committee of Quaid-e-Azam Medical college.

The data was collected from September 20,2021 to October 05,2021. Convenient sampling was done in which participants were recruited through different social media websites and mobile apps (facebook, whatsapp, telegram). Participants that met the eligibility criteria were also recruited in person by a team of medical students. The eligible data was limited to residents of Lahore, Pakistan, aged 18 and/ or above years old who have lost a loved one due to COVID-19. Participants that identified to have a previously diagnosed mental health disorder (e.g. Major depression, anxiety, schizophrenia, others) were excluded.

OpenEpi version 3 was used to calculate the sample size. Using the equation [Sample size *n* = [DEFF*Np(1-p)]/ [(d^2^/Z^2^_1-α/2_*(N-1)+p*(1-p)]], anticipated frequency of 10%, total population of Lahore, Pakistan according to the latest census 13095166, a sample size of 546 was statistically significant with 99.99% confidence level. (139 for 95%).

A sample of 808 participants submitted the survey, 71 of whom were excluded from the analysis due to age less than 18 (n=6), residents of city other than Lahore, Pakistan (n=14), reported diagnosis of mental health disorder (n=42) and patterned responses (n=9). Therefore, data of 737 participants was analyzed.

The survey questionnaire consisted of sociodemographic data and measures of prolonged grief (PG-13 scale), anxiety/depression (patient health questionnaire PHQ-4) and psychological distress (Kessler K-6) as dependent variables.

### Sociodemographic Data

The demographic data included age, gender, city of residence, marital status, employment, family income per month, education and religious belief. Loss related factors included relationship with deceased (i.e, partner, parent, grandparent, relative, friend, other relationship) and place of death (i.e home or hospital/clinic).

### Prolonged Grief Disorder (PG-13 scale)

The prolonged grief disorder scale PG-13 (23) was used to assess (PGD) prolonged grief according to ICD-11. The PG-13 instrument contains 13 items: two items (items 3 and 13) on duration and impairment that are to be answered “yes” or “no”, and 11 items assessing cognitive, behavioral and emotional symptoms, rated on a 5-point scale. Items 1, 2, 4 and 5 are rated on a frequency scale ranging from: “not at all” to “several times a day” (scoring 1–5), and items 6–12 are rated on an intensity scale ranging from “not at all” to “overwhelmingly” (scoring 1–5). To assess the presence of PGD, five criteria (A–E) must be met. These criteria include

A. an event criterion: the respondent has experienced loss;
B. a separation distress criterion: the respondent must experience grief related yearning at least daily;
C. a duration criterion: the symptoms of separation distress must be elevated at least 6 months after the loss;
D. cognitive, emotional and behavioral symptoms: the respondent must experience at least 5 symptoms based on items 4–11 at least “once a day” or “quite often” and
E. an impairment criterion: the respondent must have significant impairment in social, occupational or other important areas of functioning.

PG-13 can be scored as a continuous measure by summing the symptom items and excluding the two duration and functional impairment items. These total scores, ranging from 11 to 55 are used in the current study. There are no officially recommended cut-off scores.

### Patient Health Questionnaire-4 (PHQ-4)

PHQ-4 was used to screen for depression and anxiety (24). The 4-question scale is rated on an intensity scale ranging from ‘not at all” to ‘nearly every day”.

It ranged from total score of 0-12 with

Anxiety subscale= sum of items 1 and 2 (score range, 0 to 6)

Depression subscale= sum of items 3 and 4 (score range, 0 to 6)

On each subscale, a score of 3 or greater is considered to be positive.

### Kessler-6 (K-6)

K-6 (25) was used as a global measure of distress drawing from depressive and anxiety related symptomatology. It measures distress over a period of four weeks prior to administration of the test. It has 6 items, rated on a 5-point scale ranging from “none of the time” to “all of the time”. A total of 0-24 calibration is done and a total of 13+ is considered the cut-off point for Severe Mental Illness.

#### Statistical Analysis

The data were analyzed using the SPSS Software package (Version 26, IBM Corp., Armonk, NY, USA). The data was screened for errors and duplicates among categorical variables. The survey questions were compulsory, hence there was no missing data. Statistical weights were applied to reduce voluntary response bias. Tests of normality were applied for continuous variables to determine distribution of data. Prevalence of Prolonged Grief Disorder was assessed using the 5-point criteria of PG-13 scale. Descriptive statistics were applied for categorical variables. Pearson’s correlations coefficient applied to determine relationship between Prolonged grief scores with Phq-4 ad Kessler-6 scores. Secondary analysis to determine severity of grief with relationship to deceased and place of death was conducted using non parametric Mann Whitney-U and Kruskal Wallis-H test.

## RESULTS

### Sample Characteristics

Participants were of mean age 26 years old ranging from 18 to 54 years old, a predominantly male sample (68%) accounted for the overall sample size. Most of the participants had received higher secondary education (35.4%) with strong religious beliefs (98.8). Majority of people in sample were (57.3%) married among the sample, most participants in the study were employed (45.9%),the 40-80 thousand rupees earners having a (46.9%) presence in the data, Participants who lost their close friend comprised of 31.6% of the sample followed by those who lost their mother (17.5%). Most deaths were reported by participants to be due to covid-19. The participants who experienced death of their relatives less than 6 months were 235(31.9%), 6-12 months were 226 (30.7%) and those greater than 12 months were 276 (37.4%).

### Prevalence of PGD and Symptoms

The 13 -item PGD scale showed Cronbach’s alpha = 0.862, with all items showing good inter-item correlation (0.2-0.4). A strong cronbach’s alpha shows the questionnaire was valid in the Pakistani population, since it has not been validated before in this region. A Shapiro-Wilk test indicated that score range on PGD-13 scale did not follow a normal distribution, D (737) =0.927, p<.000, hence non parametric (mann whitney u and KW kruskal wallis test) for secondary analysis was applied. The Score ranged from 12-54 with a mean PGD score of 40.26 ± 8.54. From the total bereaved sample 110 participants fulfilled the criteria for PGD caseness, thus the total prevalence of PGD in the total sample of 737 was 15%.

Summarized below are descriptive statics of the sociodemographic factors of the study population (Table 1)

**Table 1.**

| Variables |  | PGD |  | Non PGD |  |
| --- | --- | --- | --- | --- | --- |
| Gender | Female | 29 | (26.4%) | 214 | (34.1%) |
|  | Male | 81 | (73.6%) | 412 | (65.6%) |
|  | Total | 110 | (100.0%) | 627 | (100.0%) |
| age | 18 -25 | 14 | (12.7%) | 304 | (48.5%) |
|  | 25 - 35 | 15 | (13.6%) | 127 | (20.3%) |
|  | 35-45 | 40 | (36.4%) | 117 | (18.7%) |
|  | above 45 | 41 | (37.3%) | 79 | (12.6%) |
|  | Total | 110 | (100.0%) | 627 | (100.0%) |
| education | Secondary | 14 | (12.7%) | 118 | (18.8%) |
|  | Higher Secondary | 38 | (34.5%) | 223 | (35.6%) |
|  | Graduation | 44 | (40.0%) | 207 | (33.0%) |
|  | Post-Graduation | 14 | (12.7%) | 79 | (12.6%) |
|  | Total | 110 | (100.0%) | 627 | (100.0%) |
| Marital Status | Married | 53 | (48.2%) | 369 | (58.9%) |
|  | Unmarried | 50 | (45.5%) | 233 | (37.2%) |
|  | Widowed | 7 | (6.4%) | 25 | (4.0%) |
|  | Total | 110 | (100.0%) | 627 | (100.0%) |
| employment | Employed | 58 | (52.7%) | 280 | (44.7%) |
|  | self Employed | 32 | (29.1%) | 203 | (32.4%) |
|  | unemployed | 20 | (18.2%) | 144 | (23.0%) |
|  | Total | 110 | (100.0%) | 627 | (100.0%) |
| Family income per month | 40 - 80 thousand rupees | 60 | (54.5%) | 286 | (45.6%) |
|  | above 80- thousand rupees | 21 | (19.1%) | 112 | (17.9%) |
|  | below 40 thousand rupees | 29 | (26.4%) | 229 | (36.5%) |
|  | Total | 110 | (100.0%) | 627 | (100.0%) |
| Religious belief | Yes | 109 | (99.1%) | 619 | (98.7%) |
|  | No | 1 | (0.9%) | 8 | (1.3%) |
|  | Total | 110 | (100.0%) | 627 | (100.0%) |
| The place of death was | Home | 90 | (81.8%) | 396 | (63.2%) |
|  | Hospital/Clinic | 20 | (18.2%) | 231 | (36.8%) |
|  | Total | 110 | (100.0%) | 627 | (100.0%) |
| Relation with the deceased | Close friend | 45 | (40.9%) | 188 | (30.0%) |
|  | Father | 18 | (16.4%) | 93 | (14.8%) |
|  | Husband | 7 | (6.4%) | 31 | (4.9%) |
|  | Mother | 14 | (12.7%) | 115 | (18.3%) |
|  | Other relative | 4 | (3.6%) | 61 | (9.7%) |
|  | Siblings | 12 | (10.9%) | 88 | (14.0%) |
|  | Significant other /partner * | 9 | (8.2%) | 45 | (7.2%) |
|  | Wife | 1 | (0.9%) | 6 | (1.0%) |
|  | Total | 110 | (100.0%) | 627 | (100.0%) |
| cause of the death was | covid -19 | 95 | (86.4%) | 409 | (65.2%) |
|  | post covid complications | 15 | (13.6%) | 218 | (34.8%) |
|  | Total | 110 | (100.0%) | 627 | (100.0%) |

\*Significant other /partner refers to boyfriend/ girlfriend/ engaged.

### Correlates of PGD with PHQ-4 and K-6

Mean score on Patient Health Questionnaire-4 was 7.9± 2.7 out of total of 12 (n=737). Mean score on the Kessler-6 distress scale was 13±6 out of a total score of 24. (n=737)

The relationship between prolonged grief disorder (as measured by PGD-13), anxiety and depressions (measured by phq-4) and general psychological distress (as indicated by k6 scoring) was investigated using Pearson product-moment correlation coefficient. Preliminary analyses were performed to ensure no violation of assumptions of normality, linearity and homoscedasticity. In a large sample size of n>30, the violation of normality assumption doesn’t cause problems.

People with high score on grief scale (pgd-13) also have high levels of anxiety and depression(phq-4) r=.75, df (735), p<0.001.

There is a strong positive correlation of prolonged grief with psychological distress r=.64, df (737), p<0.001, with high levels of prolonged grief associated with high level of mental distress.

### Secondary Analysis

#### Severity of Grief and place of death

The level of grief severity was compared among the places of death of the deceased. A Mann-Whitney U test revealed significant difference in the pgd score levels of deaths at home (Md = 40, n=486) and hospital (Md= 37n=251), U-42454, z=-6.7, p= (<0.01) r= 0.25 (showing medium effect size). People who lost their loved ones at homes had higher grief severity than those who lost their loved one in a hospital setting.

#### Severity of Grief and relationship with deceased

The severity of prolonged grief among the bereaved was assessed with relationships with the deceased using Kruskal Wallis test. A Kruskal-Wallis Test revealed a statistically significant difference in severity of prolonged grief among the bereaved depending on relationship with the deceased. (Gp1: n=233 close friend, Gp2: n=111 father, Gp3: n=38 husband, Gp4: n=129 mother, Gp5: n=65 other relative, Gp6: n=100 siblings, Gp7: n=54 significant other/fiancé, Gp8: n=7 wife), x2 **(df=7, n= 737) 30.9, p = 0.00**.

People who lost their wives reported the highest median score (Md=42), followed by close friends, husbands (Md=40), and other groups (Md<38).

## Discussion

As reported by the World Health Organization (WHO), COVID-19 has claimed more than 6.2 million lives globally. It is important to realize that the pandemic not only affected physical health of individuals but also compromised psychological wellbeing of both the patient and their families (26). Owing to complete isolation of patients in their final days, relatives did not get to see their loved ones for the one last time. There was also the lack of social emotional and physical support as the grieving families were unable to perform the last rites, arrange funerals and meet each other due to the worldwide lockdown. All these circumstances hyperbolized PGD in mourning individuals and made it one of the important health concerns (27).

The prevalence of PGD due to the loss of a loved one to the pandemic is roughly 3:20 of our population. Among the grieving individuals, there were more males than females. People were found to be more emotionally traumatized and experienced prolonged mourning after death at home than they were after a death in a hospital. To the author’s knowledge, this is the first cross-sectional study, conducted in Pakistan, that explores the PGD prevalence during the pandemic.

In our study, the estimated prevalence of PGD among the people who lost their loved ones to COVID-19 was estimated to be 15.4%. The prevalence of PGD in our population was much lesser than what has been reported in the Chinese population of grieving adults (37.8%) (28). This may be attributed to the fact that during the initial period of the pandemic, China was more severely affected than any other country as, with the primary case of COVID-19 being reported here, the country may have been underprepared to deal with a new virus leading to greater uncertainty and psychological repercussions among the citizens.

One of our key findings is that people who lose a loved one at home are more likely to suffer from grief disorders than people who lose a loved one in a hospital. As deaths in a hospital-based setting come with greater mental preparedness for death and awareness of complications. Prior to a loss, awareness and preparation may assist in coping with depression and accepting it (29). Previous retrospective studies have correlated low preparedness with chronic grief in caregivers after the loss of their terminally ill patients (30, 31). Additionally, a cross-sectional online survey among the bereaved in China during the COVID-19 pandemic demonstrates that mental readiness served as a protective factor towards loss (28).

Through our analysis, we observed a higher level of grief in individuals owing to the loss of their partner in comparison to other family members and friends. Male respondents were more prone to prolonged post-loss depression after losing their wives to the Covid pandemic. This is, however, in contrast to the masculine norm of more emotional control and mental stability than women, particularly in Pakistan. Assessing the ambivalence in the connection is crucial since both closeness and conflict with the deceased are positively connected with grief severity. The quality of the relationship between the bereaved and the deceased also influences the degree of grief (32).

The results also show that people with PGD are more likely to suffer from anxiety, depression and other serious mental disorders (16, 17). A latent class analysis on the Dutch population et al. (8) also identified depressive disorders in different classes of people with varying PGD symptoms. There is a likelihood that members of the primary PGD symptoms and the one with clear symptoms will likely meet the requirements for PTSD, and elevated depression during follow-ups (8).

High-risk adults should be identified early for prolonged grieving through detailed evaluation and other interventions. Immediate professional help and psychiatric treatment should be provided to grieving people who are at risk of PGD as it may lead to other mental health complications. Strong policymaking is required to include prolonged grieving and altered grief disorders in psychiatric evaluations. It is important to undertake legislative measures, such as public education campaigns and supervision of high-risk populations. It is advised that medical personnel, such as doctors, nurses, and medical students, acquire education regarding changed mourning reactions. To lessen the burden of grieving by offering evidence-based care, mental health professionals must demonstrate leadership and professionalism. Among many interventions available for psychiatric evaluation, cognitive behavioral therapy (CBT) has shown promising results in the treatment of PGD and pharmacological trials with naltrexone are being conducted (33, 34).

An important aspect to highlight here is that these psychiatric interventions come at a cost which may not be affordable by individuals belonging to low- and low-middle income countries. In regions, where focus is primarily on physical ill health, mental health needs assume secondary importance. Reaching out for help is further aggravated by the stigma that surrounds the subject of mental health in this part of the world, thus contributing to the severity of symptoms of grief and other mental health problems. Moreover, cultural and religious beliefs may interfere with asking for help. Thus, all these factors contribute to an increase in the prevalence of PGD in LMICs and thus warrant further investigations and relevant interventions to curb high levels of mental health problems in these regions.

Although our results give a picture of the estimated prevalence of PGD during the pandemic, however, owing to the cross-sectional study design, they may not be used to assess trends. In addition to this, as our data collection was mainly through an online questionnaire, further clinical and operational assessments by trained mental health practitioners are required for definite diagnosis and estimates of PGD among the adult bereaving population of Pakistan. Moreover, as the questionnaire contained questions pertaining to the death of the relative(s) of the participant, it may have been distressing to fill the required fields as the loss of a loved one can be distressing. Since we had no access to the NCOC database, it limited us to determining the total number of COVID-19 deaths during a specific period in a city. Although our study had its limitations, we believe they have not impacted the primary outcome of our study.

## Data Availability

All data produced in the present work are contained in the manuscript

